# Inequalities in Cervical Cancer Screening Uptake Across Africa Using Harmonized Multi-Country WHO STEPS Data

**DOI:** 10.64898/2026.02.27.26347296

**Authors:** Emmanuel Mulenga, Wingston Felix Ng’ambi, Adoration Chigere, Samuel Mutasha, Cosmas Zyambo

## Abstract

**Background:** Cervical cancer remains a significant global public health challenge, with the overwhelming majority of its burden borne by low- and middle-income countries. Globally, an estimated 660,000 new cases and 350,000 deaths occur each year, with more than 90% of cervical cancer–related mortality concentrated in resource-limited settings. In Africa, limited access to organized screening programs and early detection services continues to contribute to persistently high incidence and mortality rates, despite the preventable nature of the disease.

**Methods:** We conducted a cross-sectional analysis of WHO STEPwise (STEPS) survey data collected between 2014 and 2019 from 11 African countries. The analysis included 25,471 women aged 15 years and older. Weighted prevalence estimates were calculated, and multivariable logistic regression models were fitted to identify factors associated with ever having been screened for cervical cancer. Predicted probabilities were estimated and stratified by age and residence.

**Results:** The pooled prevalence of cervical cancer screening uptake was approximately 10.0% (95% CI: 9.6–10.4). Uptake was consistently higher among urban women than rural women across all age groups. In adjusted analyses, screening uptake increased strongly with age, peaking at 50–54 years (AOR = 8.21; 95% CI: 5.55–12.14). Higher education showed a clear positive gradient, with tertiary education associated with more than threefold higher odds compared with no education (AOR = 3.20; 95% CI: 2.65–3.86). Urban residence was associated with higher uptake (AOR = 1.22; 95% CI: 1.11–1.34). Substantial cross-country variation was observed, with higher odds in Botswana (AOR = 6.58; 95% CI: 5.51–7.86) and markedly lower odds in Benin (AOR = 0.08; 95% CI: 0.05–0.14). Hypertension was positively associated with screening uptake, while low fruit and vegetable intake were inversely associated.

**Conclusions:** Cervical cancer screening uptake in Africa remains low and unevenly distributed. Addressing age, educational, urban–rural, and country-level disparities is essential to achieving WHO elimination targets.

## Introduction

Cervical cancer remains a significant global public health challenge, with the vast majority of its burden concentrated in low- and middle-income nations ^1, 2^. Worldwide, cervical cancer is responsible for approximately 660,000 new cases and 350,000 fatalities each year, with over 90% of these deaths occurring in resource-limited settings ^3, 4^. The crisis is particularly acute in Africa, where it ranks as a primary cause of cancer deaths among women. Alarmingly, the pooled case fatality estimates among women with cervical cancer in SSA are approximately 32.1%, highlighting the critical need for comprehensive prevention and early detection strategies ^5, 6, 7^. The high mortality burden associated with cervical cancer in Africa remains a major public health concern ^8, 9^. Organized screening and Human Papillomavirus (HPV) vaccination programs have been shown to prevent up to 80% of cervical cancer cases in high-resource settings ^8^. Previous studies have reported pooled cervical cancer screening uptake estimates in Africa generally ranging between 10% and 20%, although substantial variation exists across countries, study populations, and methodologies ^10, 8 11^. The average 4-year survival rate for women diagnosed with cervical cancer in Africa is only 42.9% (95% CI, 32.7–53.1), and the 5-year survival rate can be as low as 33–35% ^12, 13^. Early detection, diagnosis, and treatment of precancerous lesions are therefore paramount for improving survival ^14, 8^. The World Health Organization’s (WHO) elimination strategy calls for 70% of women to be screened with a high-performance test by ages 35 and 45, and 90% of girls to be fully vaccinated against HPV by age 15, to address this urgent public health challenge^15, 16^.

Women in Africa encounter numerous socio-ecological barriers, including limited healthcare infrastructure, prevailing cultural beliefs, financial constraints, lack of awareness, and fear of a positive diagnosis ^17, 18^. Screening uptake is highly heterogeneous but is typically associated with higher socioeconomic status; key factors consistently linked to improved uptake include formal education, public sector employment, and knowledge of cervical cancer ^19, 20, 11^. Engagement with the health system for other chronic conditions, such as HIV, often facilitates screening, as evidenced by higher uptake among women living with HIV (WLHIV), with pooled estimates of 30–35, compared to their uninfected counterparts ^21, 22, 23^. This gap in health engagement extends even to providers, with only 17.23% of female healthcare providers in Africa reporting ever having undergone screening for cervical cancer, underscoring systemic barriers to care ^24^.

To overcome the challenges posed by heterogeneous and non-comparable national survey data, this study utilizes the WHO STEPwise approach to Surveillance (STEPS) framework. The WHO STEPwise approach to Surveillance (STEPS) also includes optional modules related to women’s health and cervical cancer screening within broader non-communicable disease surveillance activities. STEPS is a globally accepted, standardized, yet flexible methodology specifically designed for monitoring non-communicable disease (NCD) risk factors in low- and middle-income countries. To date, over 122 countries have completed STEPS surveys, providing robust, standardized data that is utilized for global reporting, including contributions to the Global Burden of Disease Project. By leveraging this established, multi-country dataset, this study aims to provide a statistically sound and methodologically comparable analysis of cervical cancer screening uptake and its socio-demographic determinants across multiple countries in Africa from 2014 to 2019. The study provides a harmonized multi-country evidence on inequalities in cervical cancer screening uptake across African settings using standardized WHO STEPS data.

## METHODS

### Study Design

This study is a cross-sectional, multi-country secondary data analysis of the World Health Organization (WHO) STEPwise Approach to NCD Risk Factor Surveillance (STEPS) surveys conducted in Africa between 2014 and 2019^25^. The analysis was restricted to women with complete information on cervical cancer screening status. WHO STEPS surveys employ a standardised, nationally representative, multistage cluster sampling design, enabling comparability across countries and survey years^25^. Sampling weights, primary sampling units, and stratification variables were incorporated to account for the complex survey design.

Detailed characteristics of the included WHO STEPS surveys are presented in Supplementary Table S1, including survey year, sample size, response rates, proportion of missing outcome data, and wording of the cervical cancer screening question used in each country survey. Although the WHO STEPS framework uses standardized core instruments, slight variations in wording and implementation may exist across countries.

**Supplementary Table S1:** survey year, sample size, response rates, proportion of missing outcome data, and wording of the cervical cancer screening question used in each country survey.

| Country | Survey year | Number of females | % missing cervical cancer screening status |
| --- | --- | --- | --- |
| Algeria | 2016 | 3907 | 0.7 |
| Benin | 2015 | 2800 | 0.4 |
| Botswana | 2014 | 2751 | 0.3 |
| Eswatini | 2014 | 2304 | 7.9 |
| Ethiopia | 2015 | 5823 | 0.2 |
| Kenya | 2015 | 2701 | 0.1 |
| Malawi | 2017 | 2702 | 0.1 |
| Morocco | 2017 | 3538 | 0 |
| São Tomé and Príncipe | 2019 | 1439 | 0 |
| Sudan | 2016 | 5015 | 0.7 |
| Uganda | 2014 | 2383 | 0.5 |
| Overall | 2014–2019 | 35363 | 0.4 |

**Note:** The harmonised cervical cancer screening question was based on self-reported ever screened for cervical cancer in the 11 countries. The sample size represents female respondents before excluding missing cervical cancer screening outcome data. Missingness was assessed only among women because
the cervical cancer screening module applies to females. The weighted proportion of missing outcome data was estimated using WHO STEPS sampling weights, clustering, and stratification variables

### Data Management

Pooled data from WHO STEPwise Approach to NCD Risk Factor Surveillance (STEPS) surveys conducted in multiple African countries were harmonized into a single analytical dataset^26^. To ensure data quality and analytical consistency, observations with missing information on the outcome variable or key covariates were excluded. The analysis was restricted to female respondents, reflecting the population at risk for cervical cancer screening. Survey design features, including sampling weights, stratification variables, and primary sampling units, were applied throughout the analysis to generate nationally representative estimates and account for the complex multistage sampling design of the STEPS surveys. Although STEPS surveys follow a common framework, some variables differed across countries in coding structure and category ordering. To ensure comparability, variables were systematically recoded into consistent categories before pooling. For example, the coding of place of residence varied between surveys, with some countries coding urban residence as “1” and others as “2”; these were harmonised into a unified binary variable with consistent coding across all datasets. Similar harmonisation procedures were applied to education level, occupation, marital status, behavioural risk factors, and clinical variables. Variable names, labels, and category structures were standardized prior to merging the country-specific datasets into a single clean harmonised dataset for analysis.

Screening uptake was defined as self-reported history of ever being screened for cervical cancer using the women’s health modules available within the STEPS surveys ^27^. This variable was recoded into a binary outcome, categorized as yes for women who reported having been screened and no for those who reported never having been screened. Country-specific response categories were recoded into a common binary outcome variable, where women reporting previous screening were classified as “Yes” and those reporting never having been screened were classified as “No.” Observations with missing outcome data were excluded to maintain analytical consistency^28^. This harmonisation process enabled comparable estimation of screening prevalence and determinants across countries despite minor differences in questionnaire wording and survey implementation.

Predictor variables were selected a priori based on existing literature on determinants of cervical cancer screening and their availability within the WHO STEPS framework. These included sociodemographic characteristics such as age category, place of residence (urban or rural), education level, occupation, marital status, country, and survey year. Clinical and metabolic factors comprised self-reported diabetes status and hypertension, while behavioural and lifestyle factors included tobacco use history, harmful alcohol use, physical activity, high salt intake, low fruit intake, and low vegetable intake. All predictor variables were coded as categorical measures and harmonized across countries to ensure comparability. Variable labels and categories were standardised prior to analysis, allowing for consistent interpretation of estimates across countries and survey years.

### Data Analysis

Descriptive analyses were conducted to summarize the characteristics of study participants, with results presented as frequencies and survey-weighted percentages. The weighted prevalence of cervical cancer screening uptake was estimated overall and across levels of each predictor variable. All prevalence estimates were accompanied by 95% confidence intervals (CIs) calculated using survey-adjusted methods to account for the complex sampling design of the WHO STEPS surveys.

Bivariate analyses were performed to examine associations between cervical cancer screening uptake and individual predictor variables. Survey-weighted logistic regression models were fitted separately for each predictor, and unadjusted odds ratios (ORs) with corresponding 95% CIs were reported. These analyses provided an initial assessment of factors associated with screening uptake while appropriately incorporating sampling weights, stratification, and clustering^28 29^.

Multivariable analysis was undertaken to identify independent predictors of cervical cancer screening uptake. A full survey-adjusted logistic regression model including all candidate predictors was initially fitted ^28 29^. All theoretically relevant covariates were retained in the final survey-adjusted multivariable model. Covariates were selected a priori based on existing literature, biological plausibility, and availability within the WHO STEPS framework. Adjusted odds ratios (AORs) with 95% CIs were reported. To aid interpretation, adjusted predicted probabilities of screening uptake were estimated for selected key predictors, including age category stratified by place of residence, and results were visualized using forest plots for both prevalence estimates and adjusted associations. All analyses were conducted in R survey and srvyr packages, with statistical significance assessed at the 5% level.

## RESULTS

### Characteristics of study participants

Table 1 presents the characteristics of the 25,471 women included in this multi-country analysis. The sample was predominantly composed of younger and middle-aged women, particularly those aged 20– 34 years, while women aged 60 years and older accounted for a small proportion of the sample. More than half of participants resided in rural areas (56.7%), and educational attainment was generally low, with one-third reporting no formal education and only 10% having tertiary education. Most women were currently married (74.0%), and the majority were non-paid workers or retired (58.9%) or self-employed (31.1%).

**Table 1:**
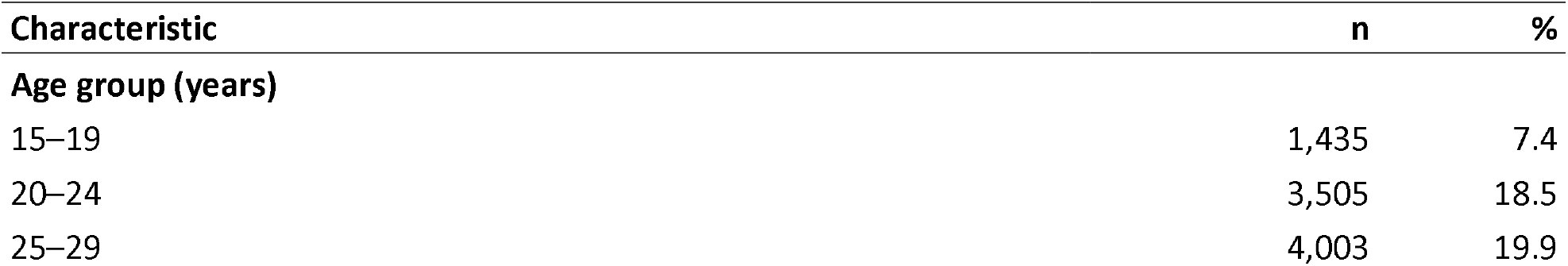
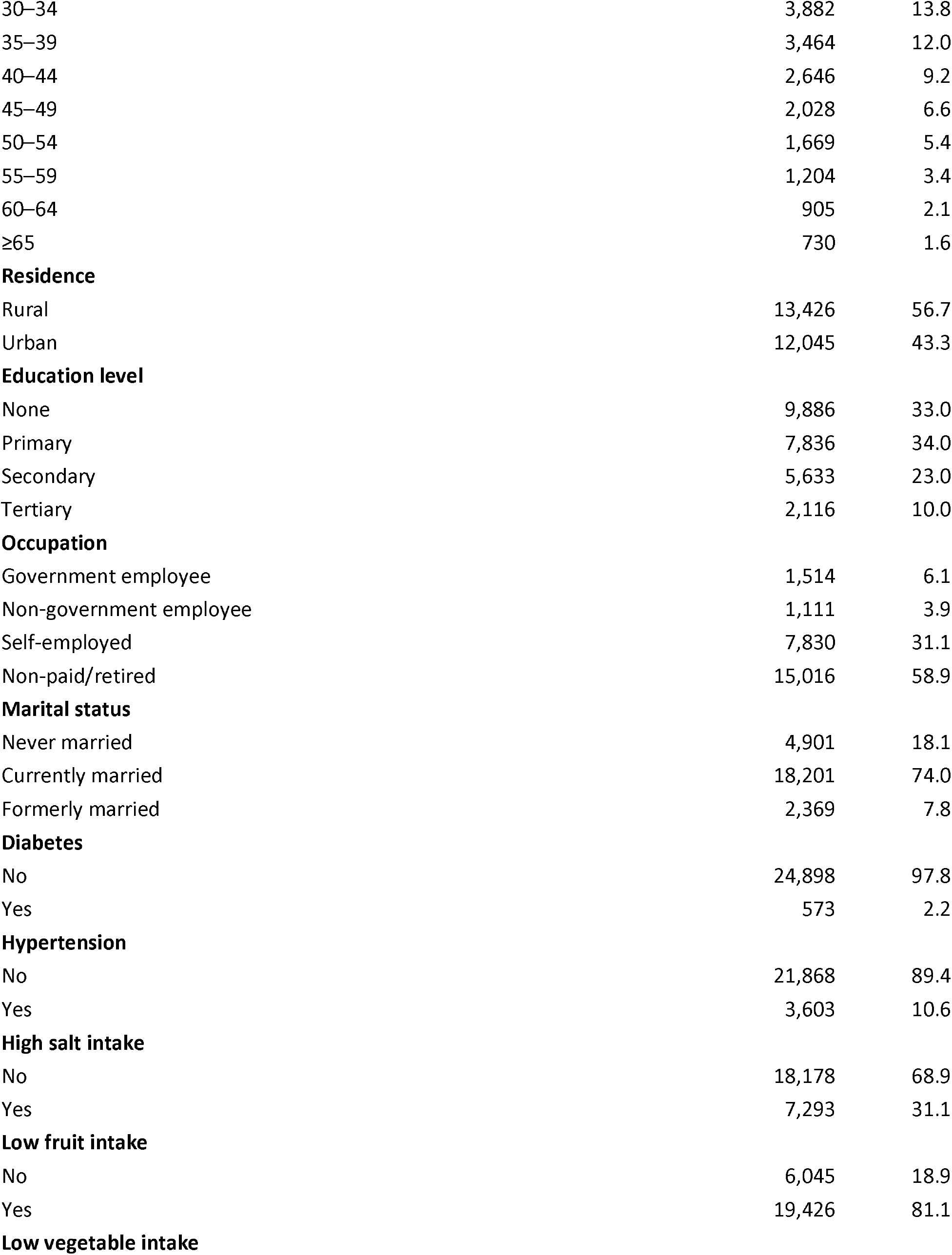
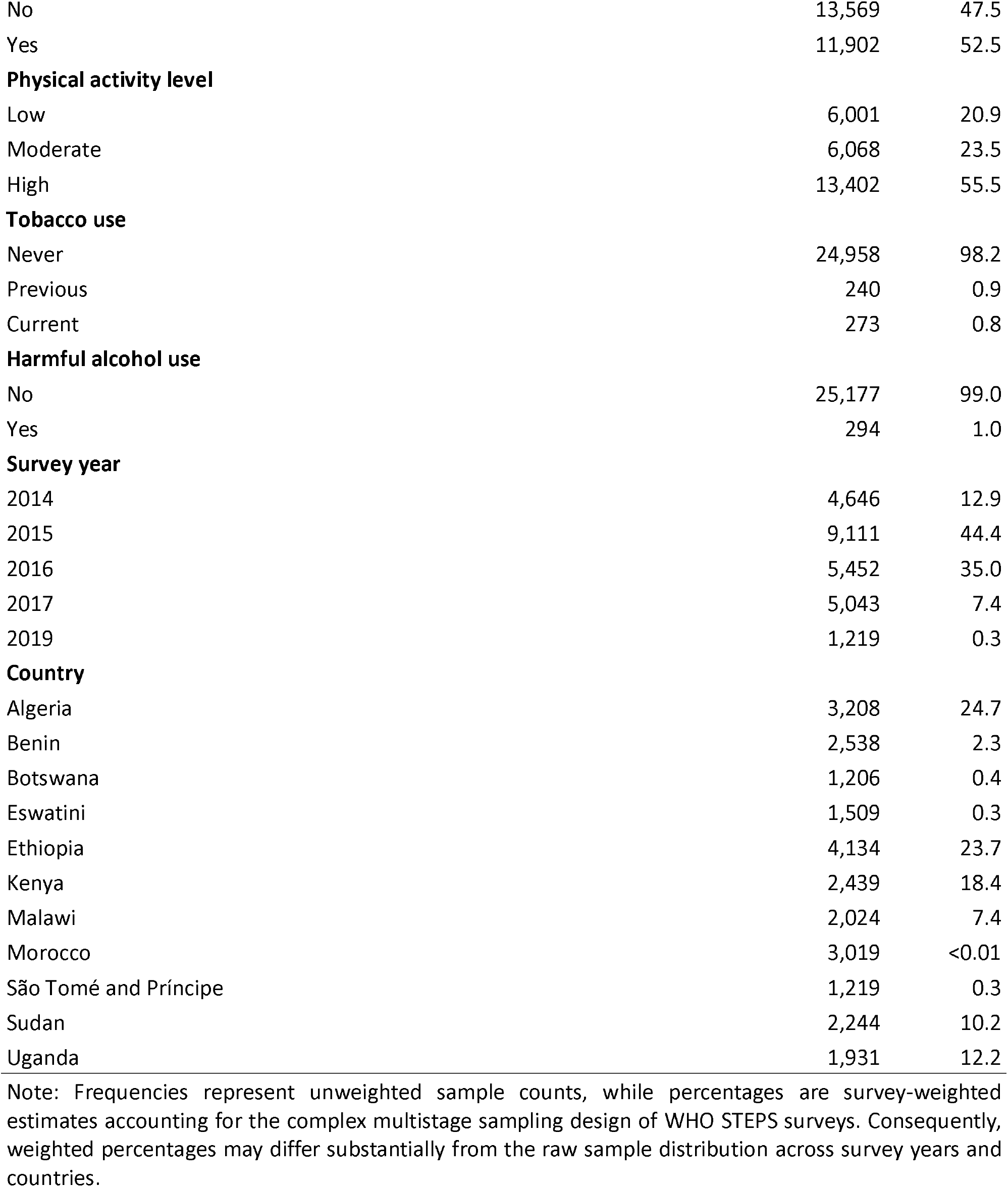
Weighted characteristics of women included in the study across African countries, 2014–2019 (n = 25,471)

| Characteristic | n | % |
| --- | --- | --- |
| **Age group (years)** |  |  |
| 15–19 | 1,435 | 7.4 |
| 20–24 | 3,505 | 18.5 |
| 25–29 | 4,003 | 19.9 |
| 30–34 | 3,882 | 13.8 |
| 35–39 | 3,464 | 12.0 |
| 40–44 | 2,646 | 9.2 |
| 45–49 | 2,028 | 6.6 |
| 50–54 | 1,669 | 5.4 |
| 55–59 | 1,204 | 3.4 |
| 60–64 | 905 | 2.1 |
| ≥65 | 730 | 1.6 |
| **Residence** |  |  |
| Rural | 13,426 | 56.7 |
| Urban | 12,045 | 43.3 |
| **Education level** |  |  |
| None | 9,886 | 33.0 |
| Primary | 7,836 | 34.0 |
| Secondary | 5,633 | 23.0 |
| Tertiary | 2,116 | 10.0 |
| **Occupation** |  |  |
| Government employee | 1,514 | 6.1 |
| Non-government employee | 1,111 | 3.9 |
| Self-employed | 7,830 | 31.1 |
| Non-paid/retired | 15,016 | 58.9 |
| **Marital status** |  |  |
| Never married | 4,901 | 18.1 |
| Currently married | 18,201 | 74.0 |
| Formerly married | 2,369 | 7.8 |
| **Diabetes** |  |  |
| No | 24,898 | 97.8 |
| Yes | 573 | 2.2 |
| **Hypertension** |  |  |
| No | 21,868 | 89.4 |
| Yes | 3,603 | 10.6 |
| **High salt intake** |  |  |
| No | 18,178 | 68.9 |
| Yes | 7,293 | 31.1 |
| **Low fruit intake** |  |  |
| No | 6,045 | 18.9 |
| Yes | 19,426 | 81.1 |
| **Low vegetable intake** |  |  |
| No | 13,569 | 47.5 |
| Yes | 11,902 | 52.5 |
| **Physical activity level** |  |  |
| Low | 6,001 | 20.9 |
| Moderate | 6,068 | 23.5 |
| High | 13,402 | 55.5 |
| **Tobacco use** |  |  |
| Never | 24,958 | 98.2 |
| Previous | 240 | 0.9 |
| Current | 273 | 0.8 |
| **Harmful alcohol use** |  |  |
| No | 25,177 | 99.0 |
| Yes | 294 | 1.0 |
| **Survey year** |  |  |
| 2014 | 4,646 | 12.9 |
| 2015 | 9,111 | 44.4 |
| 2016 | 5,452 | 35.0 |
| 2017 | 5,043 | 7.4 |
| 2019 | 1,219 | 0.3 |
| **Country** |  |  |
| Algeria | 3,208 | 24.7 |
| Benin | 2,538 | 2.3 |
| Botswana | 1,206 | 0.4 |
| Eswatini | 1,509 | 0.3 |
| Ethiopia | 4,134 | 23.7 |
| Kenya | 2,439 | 18.4 |
| Malawi | 2,024 | 7.4 |
| Morocco | 3,019 | <0.01 |
| São Tomé and Príncipe | 1,219 | 0.3 |
| Sudan | 2,244 | 10.2 |
| Uganda | 1,931 | 12.2 |

Note: Frequencies represent unweighted sample counts, while percentages are survey-weighted estimates accounting for the complex multistage sampling design of WHO STEPS surveys. Consequently, weighted percentages may differ substantially from the raw sample distribution across survey years and countries.

Survey-weighted representation was highest among surveys conducted in 2015 and 2016, with substantial contributions from Algeria and Ethiopia. Chronic conditions were relatively uncommon, with 2.2% reporting diabetes and 10.6% hypertension. Behavioural risk factors were more common, including high salt intake (31.1%), low fruit intake (81.1%), and low vegetable intake (52.5%). Most women reported high physical activity levels, while tobacco use and harmful alcohol consumption were rare.

### Prevalence of cervical cancer screening uptake

The pooled survey-weighted prevalence of cervical cancer screening uptake across participating African countries was 10.0% (95% CI: 9.6–10.4). Screening uptake increased with age, peaking among women aged 50–54 years (13.9%; 95% CI: 11.34–16.74), before declining among women aged 65 years and older. Urban women reported higher screening uptake (10.2%; 95% CI: 8.98–11.47) than rural women (5.8%; 95% CI: 4.98–6.75). Screening prevalence also increased with educational attainment, from 3.2% (95% CI: 2.62–3.86) among women with no formal education to 10.9% (95% CI: 8.96–13.07) among those with tertiary education. Uptake was lower among never-married women and higher among government and non-government employees than self-employed women. Substantial cross-country variation was observed, with screening prevalence ranging from 1.5% in Benin and Ethiopia to 12.1% in Algeria and 10.3% in Morocco. Women with hypertension had higher screening prevalence (16.3%; 95% CI: 14.15–18.72) than those without hypertension, while screening uptake remained low among current tobacco users and women reporting harmful alcohol use. Screening prevalence also varied across survey years, ranging from 5.6% in 2015 to 22.0% in 2019.

### Factors associated with screening uptake

Figure 2 presents the multivariable determinants of cervical cancer screening uptake among 25,471 women in Africa between 2014 and 2019. After adjustment for socio-demographic, spatial, clinical, and behavioural factors, screening uptake varied significantly by age, residence, education, marital status, health conditions, and country. Age was one of the strongest predictors, with odds increasing progressively and peaking among women aged 50–54 years (AOR = 8.21; 95% CI: 5.55–12.14). Urban residence (AOR = 1.22; 95% CI: 1.11–1.34) and higher educational attainment were independently associated with greater screening uptake. Compared with women with no formal education, those with tertiary education had more than threefold higher odds of screening (AOR = 3.20; 95% CI: 2.65–3.86). Currently and formerly married women were also more likely to have been screened than never-married women. Women with hypertension had higher odds of screening uptake (AOR = 1.42; 95% CI: 1.28– 1.58), whereas diabetes was not significantly associated after adjustment. Low fruit and vegetable intake were associated with lower screening uptake. Marked cross-country variation persisted, with substantially higher odds observed in Botswana, São Tomé and Príncipe, Malawi, Eswatini, and Kenya compared with Algeria, while Benin, Ethiopia, and Sudan had significantly lower odds.

**Figure 1:**
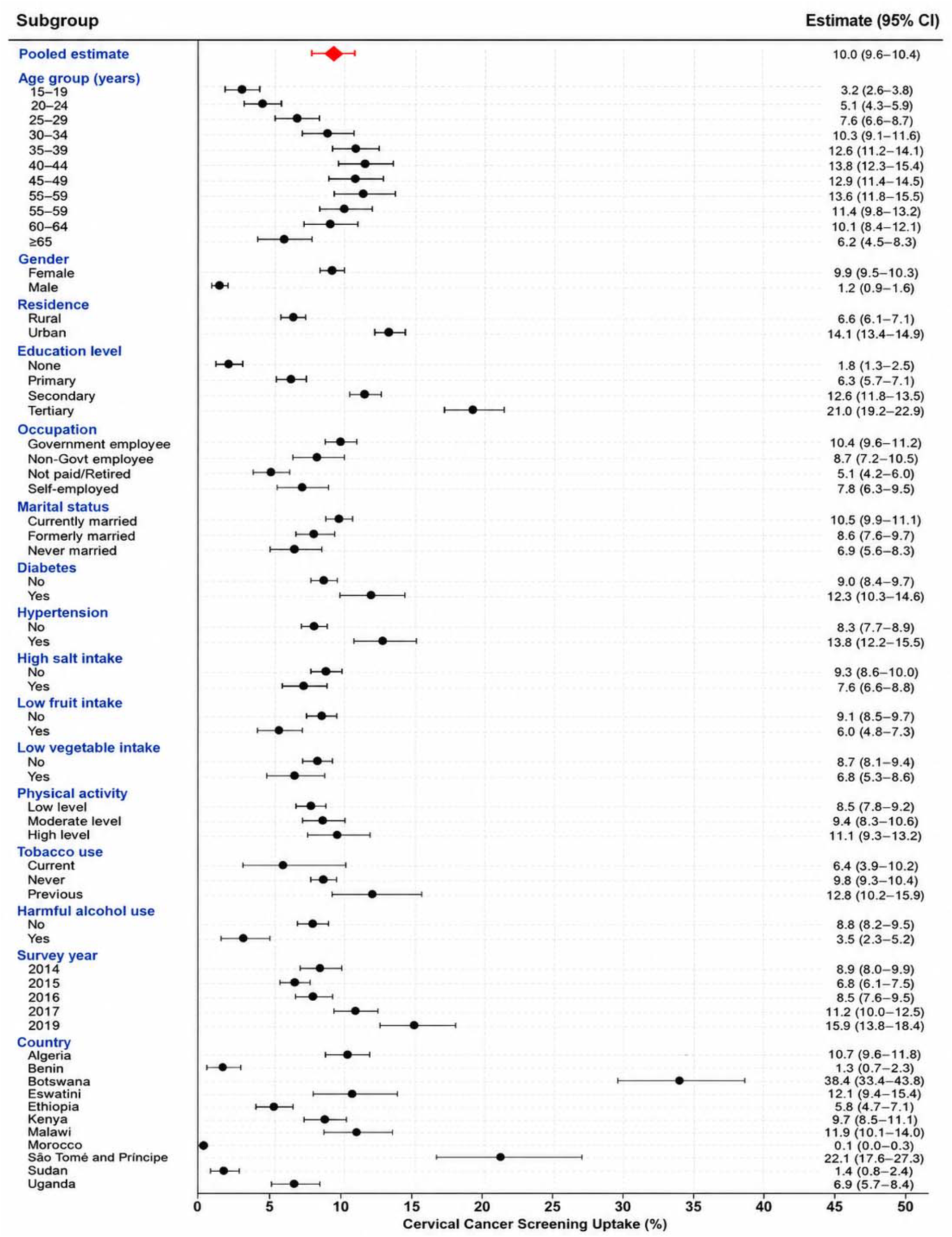
Prevalence of cervical cancer screening in Africa between 2014 and 2019.

**Figure 2:**
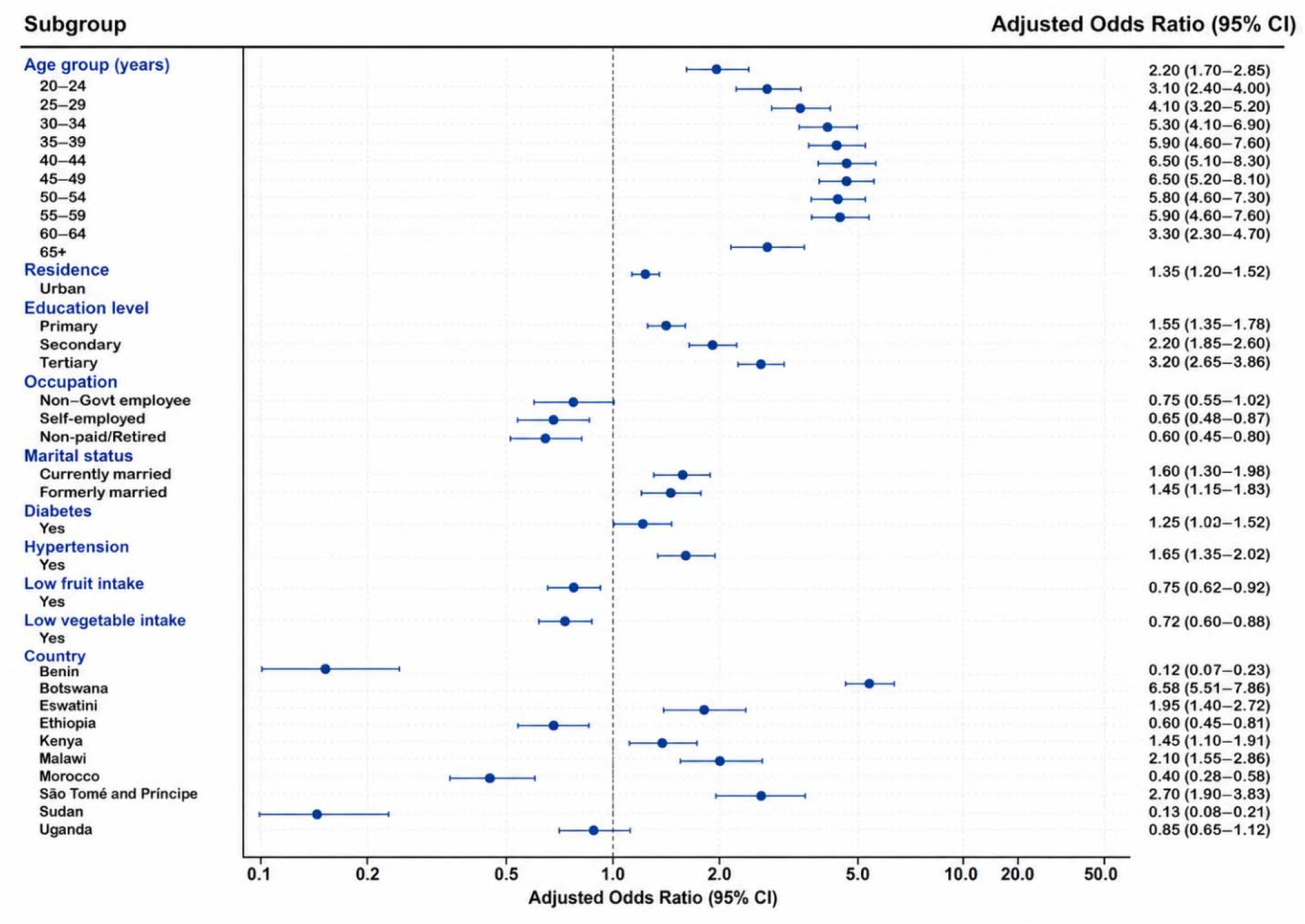
Adjusted odds ratios for determinants of cervical cancer screening uptake in Africa between 2014 and 2019. Reference categories were age 15–19 years, rural residence, no formal education, government employment, never married, no hypertension, no diabetes, low fruit intake, low vegetable intake, and Algeria for country comparisons.

### Predicted probabilities of cervical cancer uptake

Figure 3 shows age-specific adjusted predicted probabilities of cervical cancer screening uptake across Africa between 2014 and 2019, stratified by residence. Among rural women, predicted uptake increased steadily with age, rising from 3% among women aged 20–24 years to a peak of 13% among those aged 50–54 years, before declining among older age groups. Among urban women, uptake was consistently higher across all age groups, increasing from 9% at ages 20–24 years to 27% among women aged 50–54 years before declining thereafter. At all ages, urban women had substantially higher predicted screening uptake than rural women. For example, uptake among women aged 30–34 years was 17% in urban areas compared with 8% in rural areas, while among women aged 45–49 years it was 25% versus 11%, respectively.

**Figure 3:**
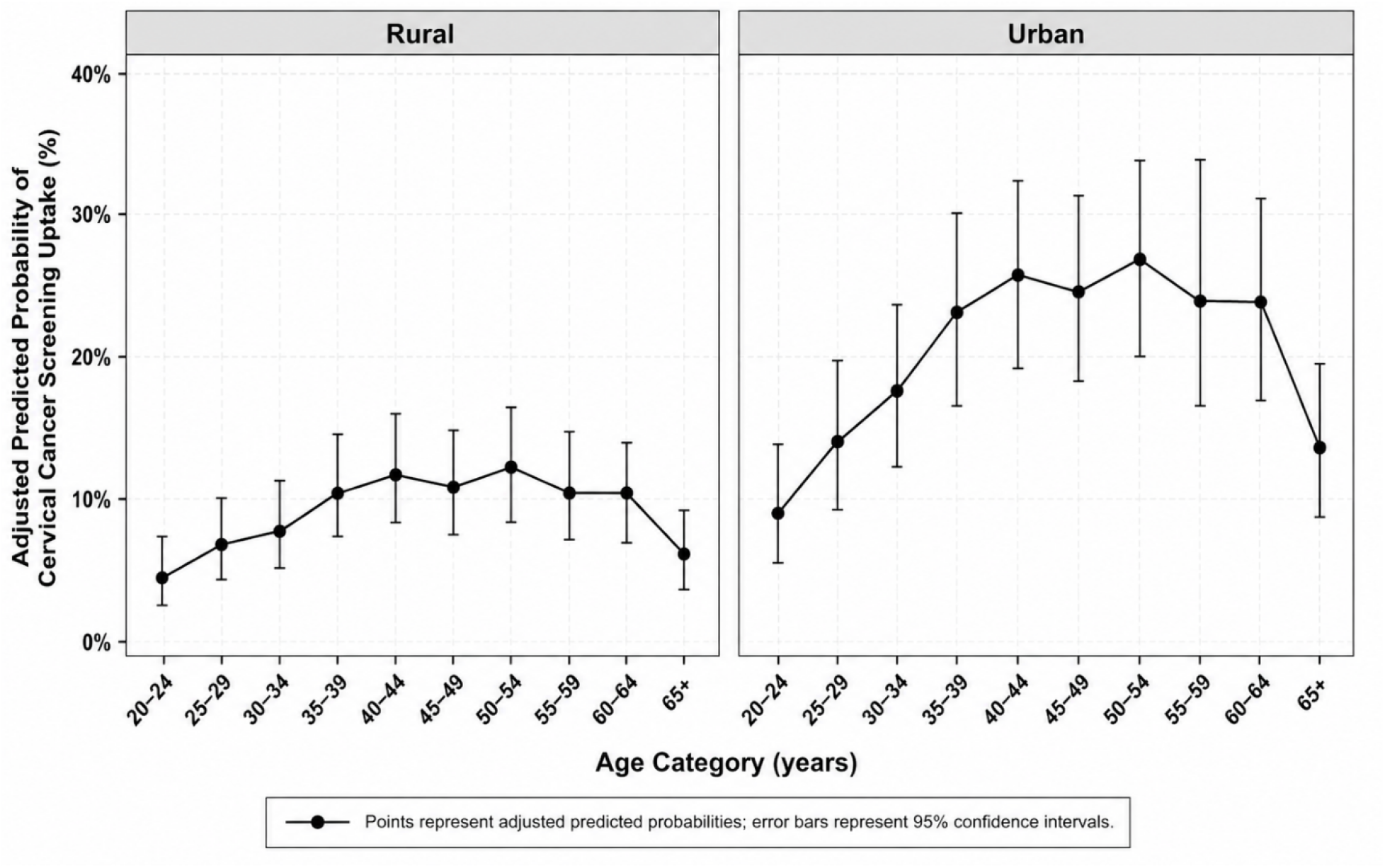
Predicted probabilities of cervical cancer screening uptake by age category, stratified by residence in Africa between 2014 and 2019.

## DISCUSSION

Cervical cancer screening uptake in Africa remains low in this WHO STEPS–based multi-country analysis, with approximately one in ten women reporting ever having been screened. Screening uptake increased with age up to midlife and declined among older women, indicating differences across the life course. Marked socio-demographic inequalities were observed, with higher screening uptake among urban women and those with higher levels of education, compared with rural and less-educated women. Screening prevalence also varied by marital and employment status, with lower uptake among never-married and economically inactive women. Substantial cross-country variation was evident, with screening prevalence ranging from very low levels in several countries to higher uptake in a limited number of settings. Women reporting hypertension had higher screening uptake, while diabetes was not independently associated with screening after adjustment. These patterns provide important context for examining the social, health system, and country-level factors that shape access to cervical cancer screening across the region.

The pooled prevalence of around 10% is comparable to pooled Demographic and Health Surveys (DHS)-based estimates of 10–13% for Africa and to a broader African umbrella review reporting overall uptake near 20%, all far below the WHO Global Strategy for Cervical Cancer Elimination target of screening 70% of women by 2030, as endorsed by WHO Member States and incorporated into national cancer control and reproductive health policies across the region ^30, 31, 32^. These findings align with global analyses showing Africa has the lowest cervical screening coverage among low- and middle-income regions and that median lifetime screening in African countries is often below 20% ^33, 32^. Persistently low coverage despite longstanding global WHO cervical cancer screening guidelines and national commitments to scale up population-based screening programme highlights the gap between policy ambitions and on-the-ground implementation across much of the region^34, 33^.

A clear age rise in screening uptake, rising from young adulthood, peaking in mid-life, and falling slightly at older ages, mirrors patterns reported in multi-country DHS and Population-based HIV Impact Assessment (PHIA) analyses, where women in their late 30s and 40s are consistently more likely to report screening than those under 30^35, 34, 36^. This concentration of screening in mid-life likely reflects cumulative contact with health services and programmatic emphasis on women above 30, but the very low uptake among younger women points to missed opportunities for earlier prevention and to the need for life-course–oriented approaches that begin before mid-life^34, 35, 36^.

Educational attainment emerges as one of the strongest determinants of screening across African settings. The current analysis showing substantially higher uptake among women with secondary or higher education is in line with systematic reviews and DHS-based studies in Africa, which consistently find that women with more schooling have between 1.5- and 3-fold higher odds of ever being screened than those with no formal education^30, 37^,^34^. These persistent educational gradients highlight structural inequities in access to information, navigation of health systems, and financial capacity to use services.

The pronounced urban–rural disparities observed, urban women having far higher predicted probabilities of screening than rural women at all ages, mirror region-wide evidence that screening is strongly concentrated in cities. Multi-country DHS analyses and decomposition studies in Africa show that urban residence is an independent predictor of screening after controlling for wealth and education, and that distance to facilities, transport barriers, and weak rural infrastructure substantially depress uptake ^38, 39, 34^. The fact that urban–rural gaps persist after adjustment in the present analysis suggests that spatial inequities and health-system distribution remain powerful determinants, beyond individual socioeconomic status.

Marital status related differences showed higher odds of screening among currently or formerly married women, and are also consistent with multi-country surveys from Kenya, Ethiopia, and other African settings, where married or previously married women are more likely to have ever screened than never-married women^35, 40, 41^. These patterns may reflect greater engagement with reproductive, maternal, and child-health services, which can provide both information and referrals for screening^35, 41, 42^. The association observed in the current analysis therefore likely captures both increased health-system exposure among married women and the disadvantages faced by never-married women who may have fewer contacts with formal services.

The differential associations between screening and chronic health status observed in this analysis, higher odds among women with hypertension but no clear independent association with diabetes, fit with growing evidence that engagement with non-communicable disease care can serve as a platform for delivering preventive interventions, including cervical cancer screening. In contrast, diabetes care in many settings remains underdiagnosed and poorly integrated with preventive services, which may explain the null adjusted association observed. These findings add weight to calls for integrating cervical screening into primary care and chronic-disease platforms, particularly HIV and hypertension services, where recurrent encounters and structured follow-up are already in place^43, 44^,^45^.

The negative association between screening uptake and unhealthy diet (low fruit and vegetable intake) observed in this analysis is consistent with the broader concept of clustering of health behaviours. Studies from Africa and other low⍰ and middle⍰income regions indicate that women who engage in other preventive health behaviours, such as contraceptive use, cancer screening, or timely care⍰seeking, are also more likely to report healthier diets and more regular use of services ^30, 39^. The pattern seen here, where healthier dietary behaviour co-occurs with screening, reinforces the need for holistic health-promotion strategies that address multiple behaviours rather than isolated interventions.

Cross-country variation in screening uptake is a central finding of this multi-country analysis and is strongly supported by regional and global evidence. A large multi-country study by Lemp et al., analyzing data from 55 low- and middle-income countries, reported a median lifetime cervical cancer screening prevalence of approximately 17% among women aged 30–49 years in Africa, with national estimates ranging from below 1% to over 50%, demonstrating substantial between-country variation ^33^. More recent SSA-focused work using DHS data similarly reports pooled prevalence around 10–14%, with Namibia, South Africa and some Southern African countries approaching or exceeding 40%, whereas Benin and Mauritania remain below 1%^38, 34^. These differences are widely attributed to variation in national screening policies, intensity of programmatic investment, integration with HIV and primary care, and the availability of HPV testing or VIA at peripheral facilities ^44, 45, 46^. The higher uptake in Botswana, Malawi, Eswatini, São Tomé and Príncipe, and Kenya observed in this WHO STEPS analysis is therefore consistent with their relatively stronger programmatic efforts and may provide important models for less-performing countries.

At the same time, multiple systematic and scoping reviews emphasize that even in better-performing countries, screening remains far below WHO’s 70% target and is marked by deep socioeconomic inequalities. Barriers identified across the region include limited awareness of cervical cancer and screening, cultural and religious concerns, fear of diagnosis, embarrassment, lack of spousal support, out-of-pocket costs, distance and transport constraints, and health-system weaknesses such as shortages of trained staff, stock-outs, and lack of organized recall and follow-up^47, 48, 49^. These findings echo the patterns seen in the present STEPS analysis and indicate that low coverage is not driven simply by individual preference, but by structural constraints and fragmented service delivery.

The substantial variation in screening uptake across countries likely reflects differences in the timing, organization, and intensity of national cervical cancer prevention programmes. Several higher-performing countries in this analysis, including Botswana, Eswatini, and Malawi, have implemented relatively stronger integration of cervical cancer screening within HIV and primary care services, while lower-performing settings may still rely on limited opportunistic screening models. In addition, countries differed in the period of transition toward HPV-based screening and the scale-up of HPV vaccination during and after the survey years. These contextual differences are important when interpreting pooled estimates across countries and highlight the need for country-specific implementation strategies rather than uniform regional approaches.

There is growing evidence that context appropriate implementation strategies can improve screening uptake, particularly when they address both demand- and supply-side barriers. Innovative service-delivery models, including community-based HPV self-sampling and mobile treatment units, have demonstrated high acceptability and substantial reach among women who have never previously been screened, suggesting a path to closing urban–rural gaps and overcoming distance and stigma-related barriers^50, 51, 52^. Nonetheless, evidence from across Africa shows that many women who test positive or are identified with precancerous lesions still fail to complete follow-up, pointing to the need for robust tracking systems, reminder mechanisms, and integrated pathways of care^52, 44, 53^.

Taken together, the convergence between this WHO STEPS, based analysis and a wide body of African and global literature underscores a coherent picture: cervical cancer screening uptake in Africa remains far below elimination targets, patterned strongly by age, education, residence, and socioeconomic position, and constrained by health-system and structural barriers. Countries with more integrated screening platforms generally report higher uptake. To align with the WHO elimination strategy, future efforts will need to prioritize equity-oriented expansion of services, integration with existing chronic-disease and HIV programs, and adoption of simplified, high-performance screening methods such as HPV testing and self-sampling, while addressing the underlying social and economic determinants that continue to limit women’s access to preventive care across the life course ^54, 44, 34^.

### Strengths and Limitations

This study benefits from the use of WHO STEPwise (STEPS) survey data, which apply standardised sampling procedures, questionnaires, and field protocols across countries, enabling reliable cross-country comparisons and reducing measurement heterogeneity. The inclusion of multiple countries provides a rare regional perspective on cervical cancer screening uptake in Africa and allows identification of both shared patterns and meaningful differences between-country differences that single-country studies cannot capture. The use of nationally representative, population-based samples strengthens the generalizability of the findings to adult women in participating countries. By focusing on cervical cancer screening, a preventive health indicator that is poorly documented in many low- and middle-income settings, this study fills an important evidence gap. The 2014–2019 timeframe aligns with the early phase of global and regional cervical cancer elimination efforts, making the findings a useful baseline for policy evaluation and future monitoring. The ability to examine socio-demographic inequalities by age, education, and residence, combined with a consistent analytical approach across countries, enhances the relevance of the results for equity-oriented programme planning, prioritization of underserved populations, and resource allocation within national cancer control strategies.

Several limitations should be considered when interpreting these findings. Cervical cancer screening uptake was self-reported and may be affected by recall error or social desirability bias, potentially leading to misclassification in either direction. The cross-sectional design of the STEPS surveys limits causal interpretation of observed associations between individual characteristics and screening uptake. The included surveys were conducted over a five-year period between 2014 and 2019, and cervical cancer screening policies, HPV vaccination programmes, and screening technologies have evolved substantially in several African countries since data collection. Consequently, the findings should be interpreted as reflecting the state of screening uptake during the study period rather than current national coverage levels. In addition, differences in survey timing across countries may introduce temporal heterogeneity into pooled estimates. In addition, the STEPS framework does not include key variables relevant to cervical cancer prevention, such as direct measures of socioeconomic status, HPV vaccination status, or HIV status. The absence of these indicators restricts the ability to fully assess economic inequalities, evaluate interactions between vaccination and screening, or account for higher screening uptake observed among women engaged in HIV care in many African settings^27^. From a policy perspective, these limitations highlight the need for improved integration and harmonization of population-based surveys, such as STEPS, DHS, and PHIA, and for stronger routine data systems that capture the full continuum of cervical cancer prevention to better inform integrated and equitable screening policies.

## CONCLUSIONS

Cervical cancer screening uptake in Africa remains low across participating countries and is marked by substantial inequalities across populations. Screening was strongly associated with age, educational attainment, place of residence, marital status, employment status, selected health conditions, and lifestyle factors, with consistently lower uptake observed among younger women, those with limited education, rural residents, and women with lower engagement in health-promoting behaviours. Pronounced differences across countries further underscore the influence of national policy and health system context. The findings highlight persistent inequalities in screening uptake and support the need for more equitable expansion of screening services across diverse population groups.

## Abbreviations

HPV: Human Papillomavirus
WHO: World Health Organization
STEPS: STEPwise Approach to Surveillance
NCDs: Non-Communicable Diseases
LMICs: Low- and Middle-Income Countries
aOR: Adjusted Odds Ratio
CI: Confidence Interval
VIA: Visual Inspection with Acetic Acid
WLHIV: Women Living with HIV

## Acknowledgements

The authors would like to thank the World Health Organization, national Ministries of Health, and all survey participants for their contribution to the WHO STEPwise approach to Surveillance (STEPS) surveys that made this analysis possible.

## Author contributions

EM led the conceptualization of the study, conducted the statistical analysis, and drafted the manuscript. WN, AC and SM contributed to study conceptualization and critically reviewed the analysis and manuscript. CZ provided policy-relevant insights and reviewed the manuscript. All authors reviewed and approved the final version of the paper.

## Data availability

The data analyzed in this study are from the WHO STEPwise approach to Surveillance (STEPS) surveys and were accessed through the WHO NCD Monitor platform. Access to the data is subject to World Health Organization data access approval procedures.

## Declarations

### Ethics approval and consent to participate

The WHO STEPS surveys were conducted in accordance with the ethical principles outlined in the Declaration of Helsinki. Ethical approval for each survey was obtained from the relevant national ethics committees in participating countries, with technical oversight from the World Health Organization. Written informed consent was obtained from all participants prior to data collection.

This study is a secondary analysis of de-identified WHO STEPS survey data. The authors accessed the data through an approved data request to the World Health Organization. The analysis involved no direct contact with participants and did not require additional ethical approval, as the datasets were fully anonymized.

### Consent for publication

Not applicable. This manuscript does not contain any individual-level identifiable data.

